# What are the best indicators of myoelectric manifestation of fatigue?

**DOI:** 10.1101/2023.03.02.23286583

**Authors:** Elvige Ornella Fegni Ndam, Étienne Goubault, Béatrice Moyen-Sylvestre, Julie N. Côté, Jason Bouffard, Fabien Dal Maso

**Affiliations:** School of Kinesiology and Physical Activity Science, Université de Montreal, Montreal, QC, Canada; Centre Interdisciplinaire de Recherche sur le Cerveau et l’Apprentissage, Montreal, Canada; Department of Kinesiology and Physical Education, McGill University, Montreal, Canada; Department of Kinesiology, Université Laval, Quebec, Canada

**Keywords:** Electromyography, Repetitive pointing task, Perception of effort, Time-frequency analysis, Complexity, Non-linear analysis

## Abstract

The myoelectric manifestation of fatigue (MMF) is predominantly assessed using median frequency and amplitude of electromyographic (EMG) signals. However, EMG has complex features so that fractals, correlation, entropy, and chaos MMF indicators were introduced to detect alteration of EMG features caused by muscle fatigue that may not be detected by linear indicators. The aim of this study was to determine the best MMF indicators. Twenty-four participants were equipped with EMG sensors on 9 shoulder muscles and performed a repetitive pointing task. They reported their rate of perceived exertion every 30 seconds and were stopped when they reached 8 or higher on the CR10 Borg scale. Partial least square regression was used to predict perceived exertion through 15 MMF indicators. In addition, the proportion of participants with a significant change between task initiation and termination was determined for each MMF indicator and muscle. The PLSR model explained 73% of the perceived exertion variance. Median frequency, mobility, spectral entropy, fuzzy entropy, and Higuchi fractal dimension had the greatest importance to predict perceived exertion and changed for 83.5% participants on average between task initiation and termination for the anterior and medial deltoids. The amplitude, activity, approximate, sample, and multiscale entropy, degree of multifractality, percent determinism and recurrent, correlation dimension, and largest Lyapunov exponent analysis MMF indicators were not efficient to assess MMF. Mobility, spectral entropy, fuzzy entropy, and Higuchi fractal dimension should be further considered to assess muscle fatigue and their combination with median frequency may further improve the assessment of muscle fatigue.

## 1 Introduction

Muscle fatigue is defined as a transient decrease in the capacity to perform physical actions [1]. Since muscle fatigue is not a physical variable by itself, its assessment requires the identification of indicators based on measurable physical variables such as force, kinematics, or electromyography (EMG) [2–4]. Myoelectric manifestations of fatigue (MMF) [5,6], measured from EMG signals, have numerous advantages such as non-invasiveness, applicability in situ [7], real-time monitoring [8], and ability to monitor fatigue of a particular muscle [9]. Considering that wearable sensors are now common in the workplace [10–12], high standard of data processing is required to extract accurate MMF information from EMG signal and better prevent work-related musculoskeletal disorders due to muscle fatigue [11,13].

A consensual approach to assess MMF is the Joint Analysis of Spectrum and Amplitude (JASA) method [14]. As the name would suggest, JASA is a computational technique that accounts for both the spectrum and amplitude of EMG signal. According to this method, muscle fatigue is characterized by a power spectrum shift toward lower frequency and amplitude increase of EMG signals. The shift of power spectrum toward lower frequency, measured via the median frequency [15,16], is attributable to the increase in motor unit synchronization [17,18] and the decrease of muscle fibers conduction velocity [6,17] possibly triggered by altered distribution of H+ and K+ ions across the sarcolemma [19] and muscle membrane excitability that increase the duration of intracellular action potentials [20]. Regarding amplitude increase, measured by the EMG activation level, it is attributable to the reduction of motor units firing rate and their increased synchronization [20,21]. However, changes in EMG activation level may not systematically reflect muscle fatigue, but rather strategies of motor unit rotation triggered by sensations related to general fatigue [22,23]. Indeed, some studies have reported a decrease of EMG activation level with fatigue during low load fatiguing manual handling tasks [24] or no change during repetitive static arm abductions [25] and repetitive work in butchers [26]. Consequently, the JASA method may provide false negative results considering that the EMG activation level does not systematically increase in the presence of muscle fatigue.

Linear indicators, such as median frequency and activation level, are based on the assumption that EMG signal is a Gaussian random process [27,28], which may have limitations to assess MMF. Indeed, EMG signals, as well as other neurophysiological signals such as electroencephalogram and electrocardiogram [29,30], have complex properties [31–34]. The complexity of the EMG signal is attributed to the mechanisms underlying its generation that contain some non-linear or chaotic features [35,36]. As a result, in recent years, several complexity-based EMG indicators have been introduced [37–41] in order to detect alteration of EMG features caused by muscle fatigue that may not be detected by linear indicators. In a recent literature review, Rampichini et al., [28] classified four groups of complexity-based MMF indicators, namely, fractal and self-similarity, correlation, entropy, and deterministic chaos. The fractal and self-similarity group includes fractal dimension [42] and degree of multifractality. During sustained isometric contractions, fractal dimension was found to decrease with fatigue [18,43–45] while the degree of multifractality increased during fatiguing static and dynamic contractions and was a more sensitive MMF than median frequency [40,46–48]. Moreover Marri and Swaminathan., [49] demonstrated that the performance of multifractal indicators were more suitable for sEMG signals as compared to monofractal fractal dimension indicators such as Higuchi fractal dimension, in their study of the classification of muscular non-fatigue and fatigue conditions using EMG and fractal algorithms. The correlation group includes correlation dimension and recurrence quantification analysis. The latter was shown to increase with muscle fatigue during biceps brachii contractions [33,50,51]. Coelho et al., [34] and Ito et al.,[52] even showed that recurrence quantification analysis was better at detecting muscle fatigue than frequency-based indicators. Although there is no evidence to date that correlation dimension is a relevant indicator of MMF, Wang et al., [28] suggested that it may be a good candidate to assess muscle fatigue. The entropy group includes sample entropy, fuzzy entropy, spectral entropy, approximate entropy, and multiscale entropy that all have showed to decrease during fatiguing isometric [53–55] and dynamic contractions [48,56]. Particularly, fuzzy and multiscale entropy were found to have a superior robustness and performance to assess MMF than frequency-based [54] and approximate and sample entropy indicator [57,58]. Finally, the chaotic properties of a non-linear system can be assessed through the largest Lyapunov exponent [59] that was shown to decrease during a fatiguing task involving the low back muscles [60]. However, as stated by Rampichini et al., [28], future standardized fatiguing protocols are needed to confirm whether the largest Lyapunov exponent could be an appropriate indicator to assess MMF. Taken together, the literature on the identification of muscle fatigue based on EMG signal has shown that complexity-based indicators seemed efficient to detect MMF. Consequently, it is now essential to determine what are the best MMF indicators to assess muscle fatigue during multi-joint movement such as repetitive fatiguing tasks.

To this end, an interesting approach is to predict the rate of perceived exertion (RPE), which is known to increase with muscle fatigue [61], from MMF indicators. Interestingly, several studies have also shown a close relationship between the RPE [62] and MMF indicators [24,63–68]. For instance, Goubault et al., [24] used correlation analyses between six MMF indicators and RPE scores assessed using the CR-10 Borg scale [69] during a laboratory simulated manual handling task. They showed that spectral entropy, median frequency, and mobility were the MMF indicators that explained the largest percentage of the RPE variance, with R-square ranging from 11% to 39%. In comparison, activity explained between 17% and 21% of the RPE variance, and activation level showed no significant relationship with the RPE [56,70,71]. In a recent study, Ni et al., [68] showed that time, frequency, and time-frequency domains MMF indicators are strongly correlated to RPE. Consequently, predicting RPE through MMF indicators is a relevant experimental paradigm to determine the best EMG-based indicators to assess muscle fatigue.

Consequently, the aim of the present study was to determine the best MMF indicators, from 15 MMF indicators identified in the literature as potentially relevant to assess muscle fatigue during a repetitive pointing task (RPT) performed with the upper limb. We hypothesized that for the anterior and medial deltoids, which are the muscles showing the largest signs of fatigue during the upper limb repetitive task used in the present studies [72–74], the mobility, median frequency, spectral entropy, fuzzy entropy, multiscale entropy [24,28,41,75,76], and degree of multifractality [40,47,49,53] will have greater importance to predict the evolution of the RPE. In addition, these MMF indicators will significantly change for a large proportion of the participants in the anterior and medial deltoid in comparison to approximate and sample entropy, recurrence quantification analysis, correlation dimension, and the largest Lyapunov exponent [28,40,46,55].

## 2 Materials and methods

### 2.1 Participants

Twenty-four right-handed participants (12 ♀; age: 32.9 ± 8.9 years old; mass: 66.8 ± 10.9 kg; height: 166 ± 9 cm) were recruited among workers exposed to repetitive tasks in a tea packaging factory. To be eligible, participants had to be free of upper-limb disabilities or musculoskeletal disorders at the time of the experiment. All participants read and signed a written informed consent form before any experimental procedure. The experimental protocol was approved by the Université de Montréal’s Ethics Committee (#CERC-19-086-D).

### 2.2 Instrumentation

#### EMG recordings

Participants were equipped with 10 wireless surface EMG electrodes (Trigno EMG Wireless System, Delsys, USA) positioned on specific anatomical landmarks of the right side of the upper limb [77,78]; namely, the anterior, medial, and posterior deltoids, the upper, middle, and lower trapezius, long head of the biceps brachii, lateral head of the triceps brachii and the serratus (Figure 1-A). Before electrode positioning, hair was shaved with a razor and skin was cleaned with alcohol swabs at the electrode sites. EMG signals were recorded at a sampling frequency of 2000 Hz.

**Figure 1.** (A) Participant equipped with EMG’s sensors (black rectangle boxes). (B) Picture of the MVIC setup. (C) Schematic top view of the RPT. Note that data collected from reflective markers are not used in the present study and results obtained from inertial measurements units sensors (orange rectangle boxes) were presented in a previous study [80].

#### Repetitive pointing task (RPT)

Two cylindrical touch-sensitive sensors (length: 6 cm, radius: 0.5 cm, Quantum Research Group Ltd, Hamble, UK) were used as proximal and distal targets for the RPT (Figure 1-B and C). The sensors were placed at shoulder height in front of the participant’s midline and at 30% (proximal target) and 100% (distal target) of the chest-arm distance. When participants touched the sensors, they delivered auditory feedback that helped participants to synchronize with the metronome as well as a transistor-transistor logic pulse recorded at 2000 Hz using Nexus software (Vicon, Oxford, UK).

#### Force

A unidirectional S-shape load cell (363-D3-300-20P3, InterTechnology Inc., Don Mills, Ontario, Canada) was used to measure the maximal voluntary isometric upward force of the right shoulder. The load cell was attached to a fixed horizontal bar located above the RPT setup (Figure 1-B).

### 2.3 Experimental protocol

Participants performed a maximal voluntary isometric contraction (MVIC) before and after the RPT described earlier. The MVIC consisted of performing a 3-sec upward maximum contraction against a fixed bar with the arm flexed at 90°. Verbal encouragement was given to participants.

The RPT consisted of alternatively pointing the proximal and distal targets with the index finger with the arm constrained to move in a horizontal plane, at a rhythm of one flexion-extension cycle per two seconds [79] with the aid of an external metronome. To this end, participants stood upright with the right arm in horizontal plane and the feet parallel at shoulder width. To ensure that participants maintained their arm horizontal, a mesh barrier was placed under the elbow trajectory. Participants’ left arm rested on the side of the body. This task has been shown to fatigue the anterior and medial deltoid [72–74]. Every 30 seconds, participants reported their RPE using the CR-10 Borg scale (RPE) [62] without interrupting the RPT movement. Additionally, every 2 minutes, the RPT was interrupted so that participants could perform a MVIC as described above. Immediately after the MVIC, participants resumed the RPT without resting. Participants were asked to “perform the task for as long as possible”. They were stopped as soon as they reached a score of 8 or higher on the RPE scale but were not aware of this stoppage criteria.

### 2.4 Data preprocessing

Data processing was performed using Matlab R2019a (The MathWorks Inc., Natick, MA, USA). EMG data were filtered using a 2^nd^ order Butterworth zero-lag 10-400 Hz bandpass filter. Data were then zero-aligned by subtracting the mean signal value and resampled to 1000 Hz to reduce computation time. For the calculation on each indicator, each participant’s data was segmented into flexion-extension cycles. Each indicator described below was calculated for the 5 cycles preceding each RPE for the PLSR analysis, and for the first 10 (RPT initiation) and last 10 (RPT termination) for the analysis of the proportion of change of MMF indicators among participants.

#### 2.4.1 Data processing: linear MMF indicators

*Median frequency* was calculated using the following formula:

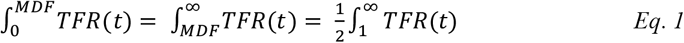

*TFR* is the power spectral density calculated in the time-frequency resolution; the time-frequency analysis was performed by applying a continuous Morlet wavelet transform (wave number: 7, frequency range: 1 to 400 Hz in 1 Hz steps) to the pre-processed EMG signals [81] (WavCrossSpec Matlab package), *t* is a time instant.

*Spectral entropy* was computed as follow [24]:

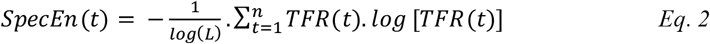

where, *L* is the number of spectral components in the EMG spectrum, *TFR* is a power spectral density calculated in time-frequency, *t* is a time instant, *n* is the number of seconds in the trial. *Activation levels* were obtained from 9 Hz low-pass filtering of the full-wave rectified EMG signals normalized by the maximum voluntary muscle activation [24] obtained using the average of the maximum 2-sec non-consecutive window across all MVIC tests.

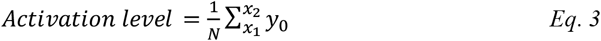

where *x*_1_ and *x*_2_ represent the muscle activation segment, *N* represents the number of elements between *x*_1_ and *x*_2_, and *y*_0_ represents the normalized EMG envelop of the signal.

*Activity* is the measure of the variance (*σ*_0_) of the signal [56,70,71].

*Mobility* is defined as the root square of the ratio between the variance of the first derivative of the signal and the variance of the signal [24,56,70,71]. The first-time derivative of the EMG signal was calculated on the entire signal using the following equation:

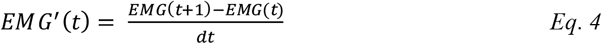

The variance of the first-time derivative of the EMG signal and the variance of the EMG signal were then calculated on each muscle activation segment, before calculating the root square of the ratio between both.

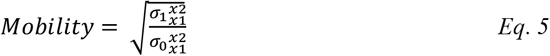

where 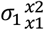 represents the variance of the first derivative of the EMG signal for muscle activation segment between *x*_1_ and *x*_2_, and 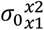 is the variance of the EMG signal for muscle activation segment between *x*_1_ and *x*_2_.

#### 2.4.2 Data processing: Non-linear MMF indicators

##### 2.4.2.1 Fractals Self-Similarity

*Higuchi fractal dimension (HFD)* was computed using the algorithm proposed by Higuchi (1988). Briefly, EMG is analyzed in time, as a sequence of samples *x(1), x(2)*,…, *x(N)*, and *k* new self-similar time series 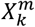 constructed as [42,82]:

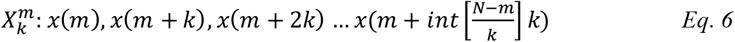

for the initial time *m = 1, 2*,…, *k*; the time interval *k = 2*,…, *k*_*max*_ [82]; and *Int[r]* the integer part of a real number *r*. Then, the length of every *L*_*m*_(*k*) is calculated for each time series or curves 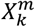 as:

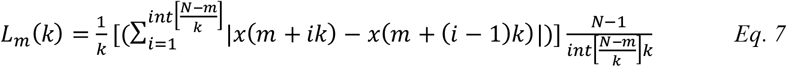

and is averaged for all *m*, therefore forming an average value of a curve length *L(k)* for each *k=2*, …, *k*_*max*_ such as:

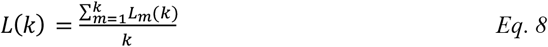

Finally, *HFD* is evaluated as the slope of the best-fit form of *ln(L(k))* vs. *ln(1/k)*:

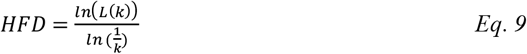

*HFD* was calculated using the Higuchi FD Matlab function [83] using *k*_*max*_ *= 8*.

*Degree of multifractality (DOM)* was calculated from the *multifractal detrended fluctuation analysis (MFDFA)* [40,47]. For an EMG signal {x(t), t = 1, 2, …, N}, *MFDF* analysis first involves a random walk construction in this form [84]:

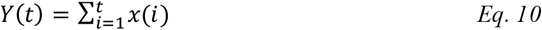

where {*Y(t), t = 1, 2*, …, *N*}. Next, a moving average function is computed in a moving window represented as:

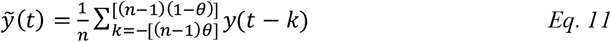

where *n* is window size, [(*n* − 1)(1 − *θ*)] is largest integer not greater than *x*, [(*n* − 1)*θ*] is smallest integer not smaller than *x*, and *θ* is a position parameter with value varying in between 0 and 1. After three more steps consisting on detrending the signal series to get the residual, then dividing the residual series into *M* disjoint segment with same size *n*, where 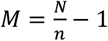, and finally determining the *q*^*th*^ order overall fluctuation function, the power law can be determined by varying the segment size *n* for fluctuation function as:

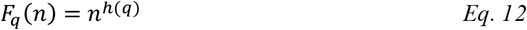

The multifractal scaling exponent is given as:

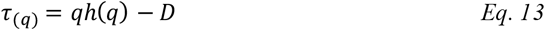

where *D* is the fractal dimension of geometric support of multifractal measure [85]. The singularity strength function and multifractal spectrum are obtained using Legendre transform [86] and represented as respectively followed:

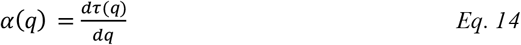

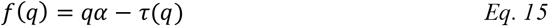

Finally, the degree of multifractality is measured as the distance between maximum exponent and minimum exponent in multifractal spectrum:

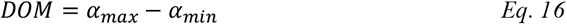

For *MFDF* computation, the MFDFA1 MATLAB function was used with a scale *q* varying between -10 and 10 with increments of 0.1, and *m* varying between 2^2^ and 2^12^ with increments of 2^0.1^ as input parameters [40,47].

##### 2.4.2.2 Entropy

*Approximate entropy* was computed as follow:

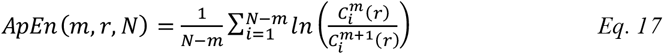

where the embedding dimension *m* = 2, the distance threshold *r* = 0.25 [58], *N* the number of sample in the time series, and 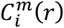 the number of vector *u(i)* within the distance *r* from the template vector *u(i)* computed as:

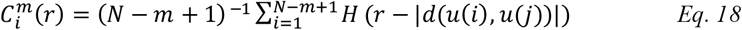

with *H ()* being the Heaviside step function where *H* is 1 if (*r* − |*d*(*u*(*i*), *u*(*j*)) ≥ 0 and 0 otherwise. For the computation, the ApEn Matlab function [87] was used with the mentioned above parameters as input.

*Sample entropy* can be defined mathematically by:

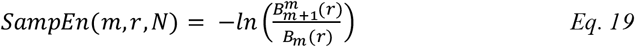

with *Bm*(*r*) defined as:

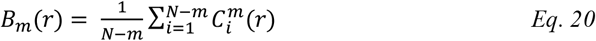

where *B*_*m*_(*r*) is the number of matches of length *m* and 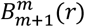 as the subset of *B*_*m*_(*r*) that also matches for length *m+1*. Here the embedding dimension *m* is equal to 2 and the tolerance *r* = 0.25 [55,56] and *N* the number of sample in the time series. For the computation, the SampEn Matlab function [88] was used with the mentioned above parameters as input.

*Fuzzy Entropy* was computed as follow [28,54,55]:

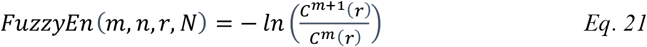

where *N* is the number of samples in the time series, *C*^*m*^(*r*) is the average of 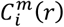 calculated as:

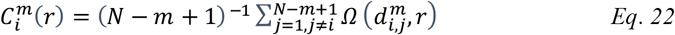

with

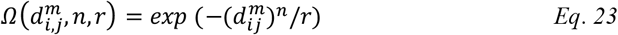

using an embedding dimension *m* = 2, a power factor *n* = 2 and a tolerance *r* = 0.25 [54,55,58]. For the computation, the FuzzyEn Matlab function [89] was used with the mentioned above parameters as input.

*Multiscale Sample Entropy* was calculated using the rapid refined composite multiscale sample entropy algorithm (R2CMSE) [53].

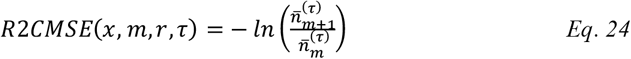

with 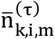 the number of vector-matching pairs defined as:

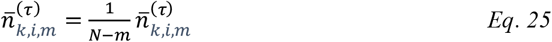

where *N* is the number of sample in the time series, the embedding dimension *m* = 2, a tolerance *r* = 0.25, and a time scale factor *τ* = 20 [53,90]. For the computation, the R2CMSE Matlab function [53] was used with the mentioned above parameters as input.

##### 2.4.2.3 Correlation

*Recurrence Quantification Analysis (RQA)* was performed using the percent of determinism method (%DET) which quantifies the amount of rule-obeying structures present within any physiological signal, and the percent of recurrence (%REC) which quantifies signal correlations in higher dimensional space [28,91].

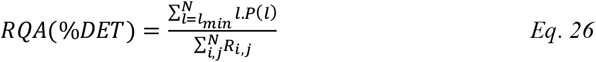

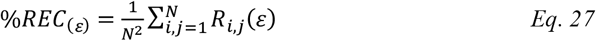

where *P(l)* is the frequency distribution of diagonal lines, with length *l* an integer number, and *R*_*i,j*_ the Euclidian distance matrix created from the phase space and transformed into a recurrence plot, using a time delay *τ = 4ms* [33,51], an embedding dimension *d = 15* [51], and a threshold value *r = 2* [92]. For the *RQA* computation, the RPplot Matlab function [92] was first used with the mentioned above parameters as input, before using the recu_RQA Matlab function with *I = 1*.

*Correlation dimension* is a measure of the amount of correlation contained in a signal connected to the fractal dimension. Its estimation requires the calculation of the correlation integral *C(r)*, which is the mean probability that the states of the dynamical systems at two different times are close, i.e., within a sphere of radius *r* in the space of the phases [28]. Given a time series *g*(*k*), the phase space is reconstructed by the vectors:

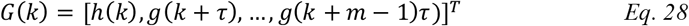

with *m* the embedding dimension and *τ* a delay. The correlation integral is then estimated by the sum:

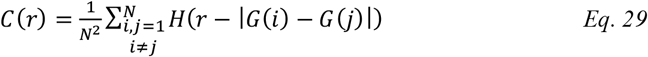

where *N* is the number of states, and *H()* the Heaviside function. If *g*(*k*) is the output of a complex system, when *N* increases and *r* decreases, *C*(*r*) tends to increase as a power of *r, C*(*r*)∼*r*^*CD*^. The correlation dimension of the system can then be estimated as the slope of the straight line of best fit in the linear scaling range region in a plot of *ln(r)* versus *ln r* [28]. For the *correlation dimension* computation, the correlationDimension Matlab function was used.

##### 2.4.2.4 Chaos

*Largest Lyapunov exponent* can be calculated using the Rosenstein’s method as follow (Rosenstein, Collins, et De Luca 1993; Chakraborty et Parbat 2017; Rampichini et al. 2020):

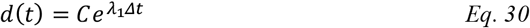

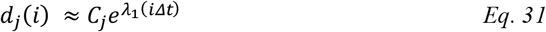

where *d(t)* is the average divergence at time *t*, and *C* is a constant that normalizes the initial separation. By taking logarithm of both sides of the equation above, we obtain:

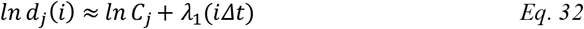

It represents a set of approximately parallel lines (for *j=1, 2*, …, *M*), each with a slope roughly proportional to *λ*_1_. The largest Lyapunov exponent is easily and accurately calculated using a least-squares fit to the “average” line defined by:

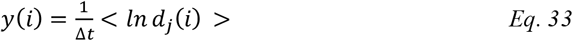

Where < > denotes the average over all values of *j*. For the *largest Lyapunov exponent* computation, the lyaprosen Matlab function [93] was used with *dt = 0.1* as input parameter.

#### 2.4.3 Statistical analyses

Firstly, a paired t-test was conducted on the maximum voluntary isometric force to assess the effect of RPT on maximum force.

Then, partial least square regression (PLSR) analyses were performed using z-score values of MMF indicators as predictors and z-score values of RPE as response variable. PLSR is particularly suited when the number of predictors is larger than the observations, and when there is multicollinearity among predictors [94,95], which is the case for predicting RPE scores from the combination of 135 input variable (9 muscles * 15 MMF indicators) in this study. To avoid overfitting of the data, the number of latent variables was determined when the absolute error of RPE prediction was minimal [94]. In order to calculate the performance of our approach, the whole data was divided into training and testing sets, and 5-folds cross-validation was used subsequently to reduce the bias due to random sampling of the training and test sets [97,98]. To do so, the whole data set was randomly split into 5 mutually exclusive subsets (folds) i.e., 4-folds included 17 participants used as the training set and 1 - fold included 4 participants used as the testing set after excluding three participants because of missing data. For each model, the absolute error of prediction was calculated on denormalized values. The cross-validation accuracy was calculated as the average of the 5 individual accuracy measures [99,100]. The variable importance in projection (VIP) was calculated for each model to determine the most relevant MMF indicators to explain variation of RPE. This whole PLSR procedure was repeated 100 times in order to have one hundred partitions of cross-validation, reducing the bias due to random sampling of the training and testing sets [101,102]. Then model performance (i.e., absolute error of prediction) was averaged over the 500 data (100 repetitions * 5-folds). Finally, an unpaired t-test was performed between the VIP values of the median frequency and the activation level and the VIP values of each MMF indicator of each muscle and the to assess the efficiency of the other indicators compared to the JASA indicators.

The effect of the RPT for each MMF indicator and muscle was assessed on a participant-specific basis using an unpaired t-tests between the 10 first cycles (RPT initiation) and the last 10 cycles (RPT termination) followed by Cohen’s *d* effect size computation. We then determined the percentage of participants for which there was a significant change, namely, a significant decrease for the median frequency, mobility, Higuchi fractal dimension, correlation dimension, largest Lyapunov exponent, and spectral, approximate, sample, multiscale, and fuzzy entropy indicators [17,18,56,103–105], and a significant increase for the activation level, activity, degree of multifractality, percent of determinism, and percent of recurrence [5,33,56,84].

## 3 Results

### 3.1 Maximum voluntary isometric force

The paired t-test revealed a significant RPT effect (p<0.001) on maximum voluntary isometric force. The MVIC performed immediately after the RPT termination was significantly smaller than the MVIC performed before the RPT initiation (Figure 2).

**Figure 2.**
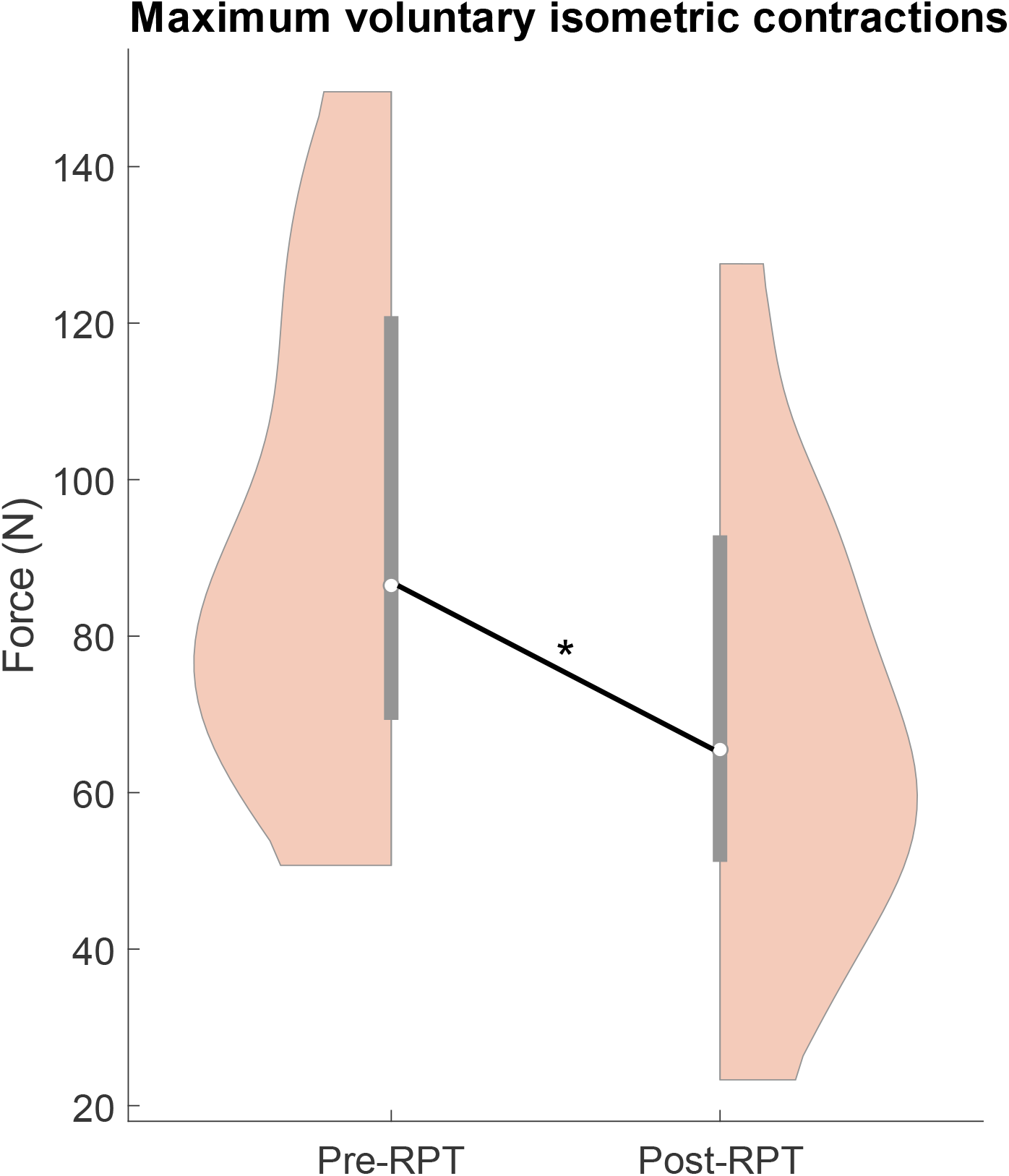
Violin plot representation of maximum voluntary isometric force during pre- and post-RPT. On each violin representation, the white dot represents the median, the thick gray bar represents the interquartile range, and the pink area violin-shaped represents the data distribution.

### 3.2 Partial least square models

#### 3.2.1 Performance of the partial least square regression to predict RPE

Over the 100 iterations, the PLSR model that generated the smallest mean absolute error of prediction of RPE values on the testing set included two latent variables (1.36 ± 0.29; Figure 3, right). This model explained 73.36 ± 2.94 % of the variance of the RPE on the training set (Figure 3, left). Although models including more latent variables explained a greater percentage of RPE variance on the training set than the two-latent variable model, the absolute error of prediction on the testing set was greater than for the two-latent variable model indicating an overfitting [106]. Consequently, the VIP results in the next section were computed from the two-latent variable model.

**Figure 3.**
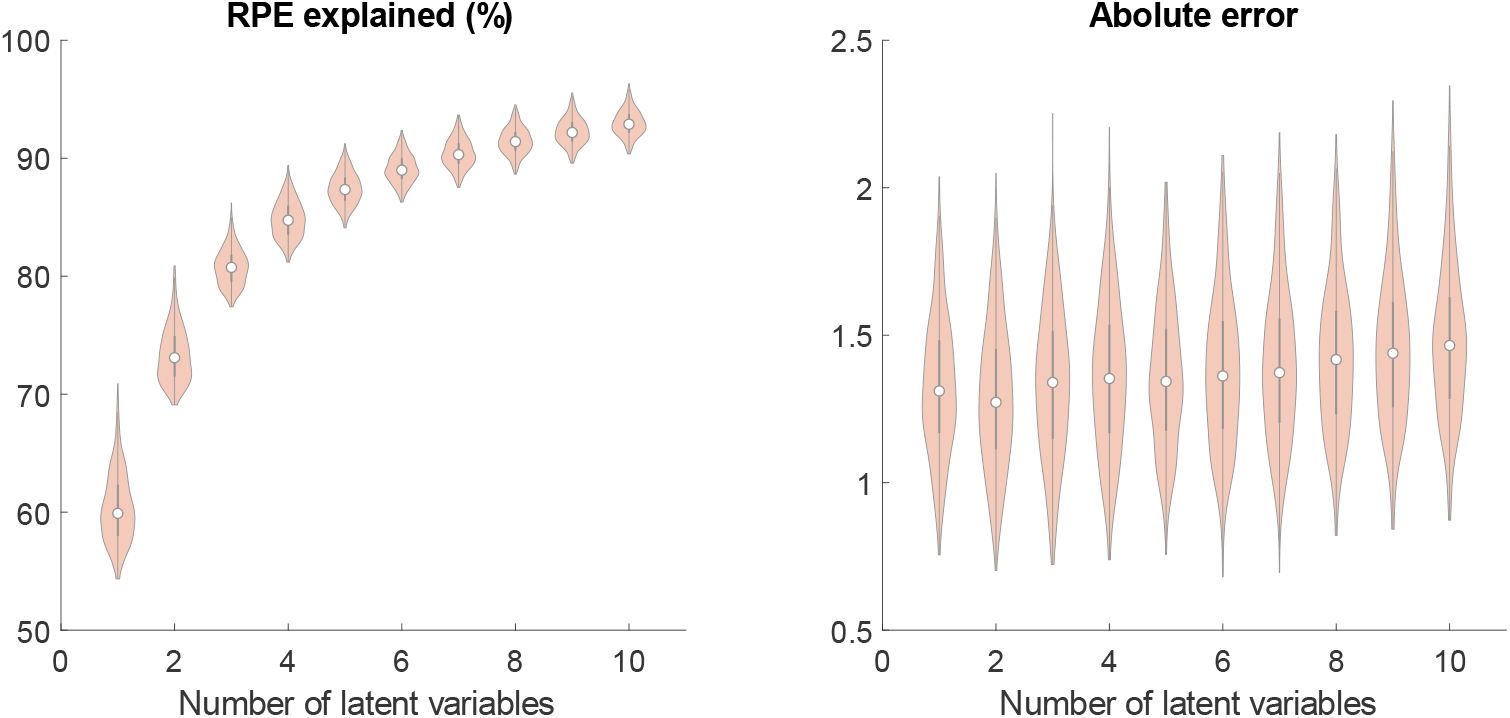
Violin plot representation of the percentage of RPE explained obtained on the training set (left) and absolute error of prediction obtained on the testing set (right) for the 100 iterations of the 5-fold cross-validation of the PLSR analysis. On each violin representation, the white dot represents the median, the thick gray bar represents the interquartile range, and the pink area violin-shaped represents the data distribution.

#### 3.2.2 VIP values for each MMF indicator and muscle

The highest VIP value was obtained for the median frequency of the medial deltoid (2.28 ± 0.11) and was significantly higher than the VIP of any other MMF indicator of this muscle (Figure 4). The VIP value of the activation level (1.41 ± 0.18) was significantly lower than the VIP value of the mobility (2.10 ± 0.14), Higuchi fractal dimension (2.03 ± 0.14), spectral entropy (1.89 ± 0.15), and fuzzy entropy (1.77 ± 0.10). All other MMF indicators had significantly lower VIP values than the VIP value of the activation level. The VIP values obtained for the anterior deltoid showed similar pattern for the median frequency (1.77 ± 0.14) that was significantly higher than the VIP value of any other MMF. Then, fractal Higuchi (1.47 ± 0.14), spectral entropy (1.41 ± 0.14) and fuzzy entropy (1.26 ± 0.12) had qualitatively higher VIP values than the activation level but did not reach significancy.

**Figure 4.**
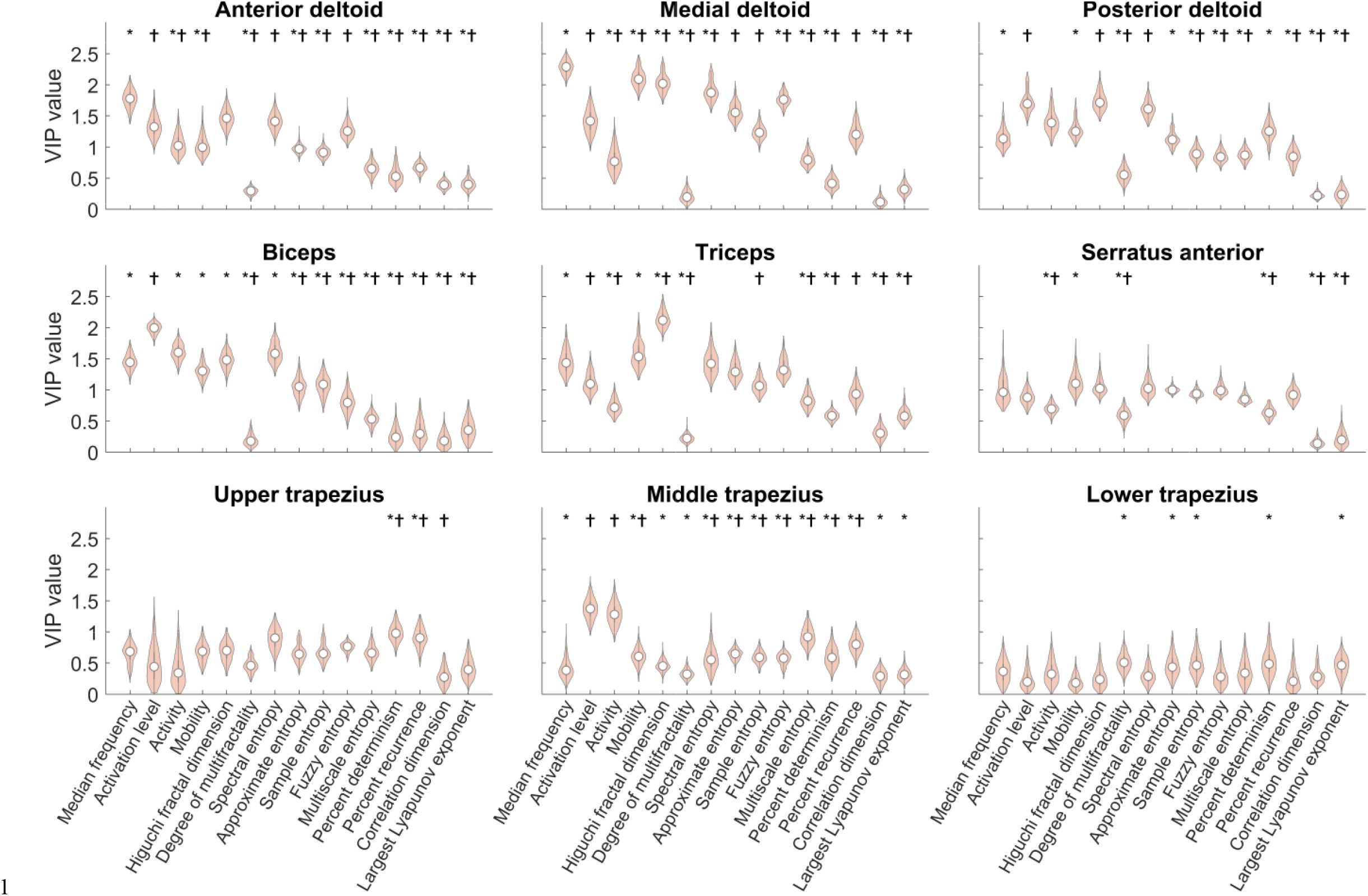
Violin plot representation of VIP values for all muscles and MMF indicators. On each violin representation, the white dot represents the median, the thick gray bar represents the interquartile range, and the pink area violin-shaped represents the data distribution. An * indicates a significant difference between activation level and a MMF indicator and a † indicates a significant difference between median frequency and a MMF indicator.

For the posterior deltoid, activation level (1.73 ± 0.17) had VIP values significantly higher than the VIP values of all indicators except Higuchi fractal dimension (1.74 ± 0.15). For the biceps, the VIP value of the median frequency (1.46 ± 0.12) was significantly smaller than the VIP value of the mobility (1.57 ± 0.13) and the Higuchi fractal dimension (2.13 ± 0.14); the VIP value of the activation level (1.12 ± 0.09) was significantly smaller than the VIP value of the mobility (1.57 ± 0.13), Higuchi fractal dimension (2.13 ± 0.14), spectral entropy (1.44 ± 0.16), approximate entropy (1.31 ± 0.15), and fuzzy entropy (1.35 ± 0.16). For the triceps, the VIP value of the activation level (1.98 ± 0.19) was significantly greater than the VIP value of activity (1.61 ± 0.12), mobility (1.31 ± 0.20), Higuchi fractal dimension (1.48 ± 0.13) and spectral entropy (1.60 ± 0.20). Finally, the MMF indicators of the serratus anterior, upper, middle, and lower trapezius had the lowest VIP values.

#### 3.2.3 Proportion of change of MMF indicators among participants

Figures in supplementary material S1-9 (one figure per muscle) show row values of each MMF indicator for each participant at RPT initiation and termination. Concerning the medial deltoid, the proportion of the participants for whom there was a change between initiation and termination for median frequency, mobility, Higuchi fractal dimension, spectral entropy, fuzzy entropy, and percent of determinism, was 90%, 80%, 90%, 86%, 90% and 81%, respectively. These indicators also showed the strongest effect sizes (Cohen’s *d* ranged between 3.5-5.35, the percent of determinism excluded, which had a Cohen’s d value equal to 2.13) (Figure 5). Strong effect sizes were also observed for approximate entropy (Cohen’s *d*=4) and activation level (Cohen’s *d*=3.6). The degree of multifractality, multiscale entropy, correlation dimension, and Lyapunov exponent changed for only 0%-45% participants with Cohen’s *d* effect sizes below 1.1. Concerning the anterior deltoid, the median frequency, activation level, activity, mobility, Higuchi fractal dimension, spectral entropy and fuzzy entropy were the MMF indicators that also showed significant change between RPT initiation and termination for a large proportion of the participants, i.e., 85%, 76%, 74%, 80%, 80%, 84%, 65% (Cohens’ *d* effect sizes ranged between 2.05-3.16).

**Figure 5.**
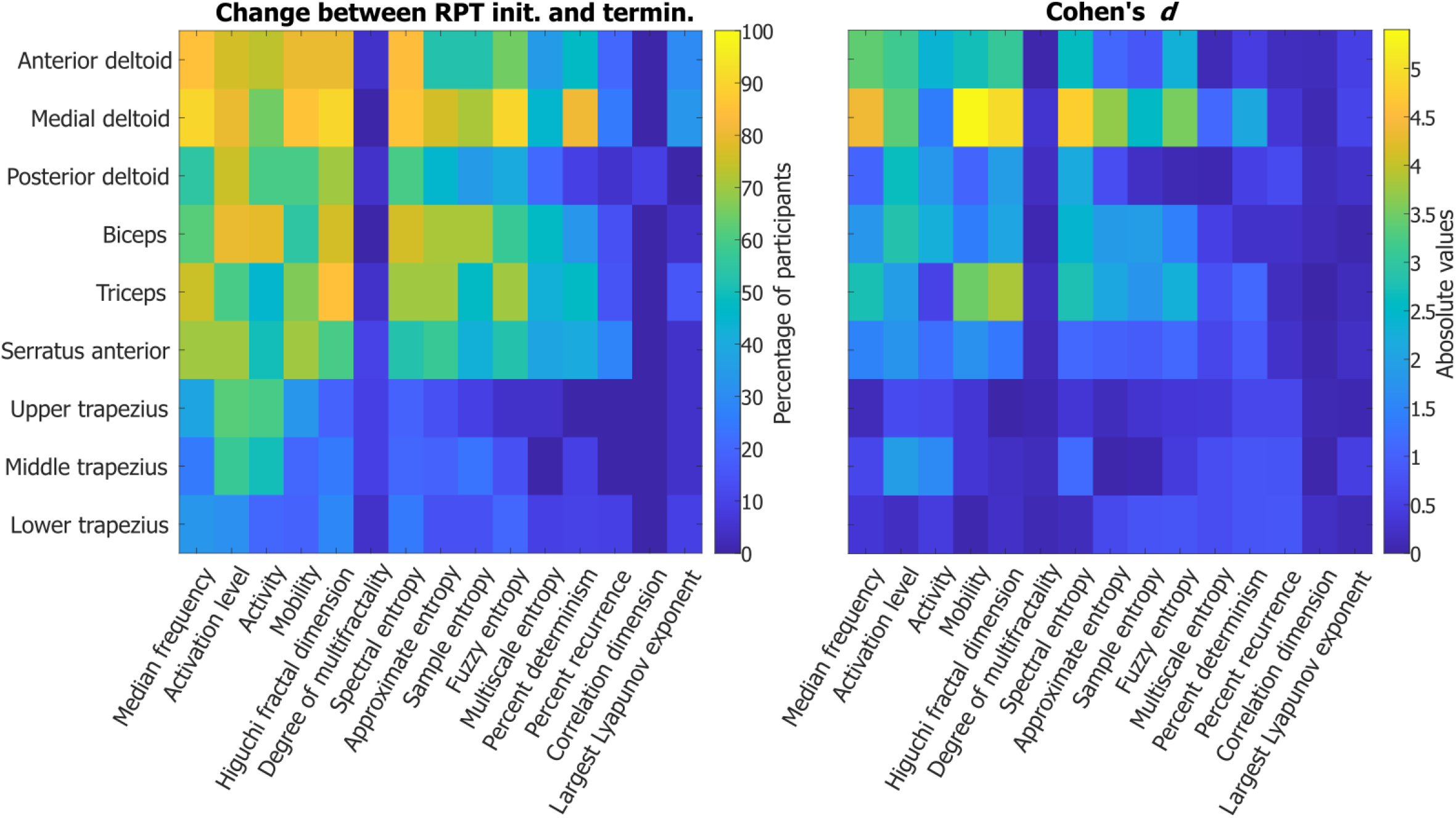
Heat maps representation of the percentage of participants showing significant effect of time between RPT initiation and termination on unpaired t-test (left column) and their corresponding Cohen’s *d* effect sizes (right column) for each muscle and each MMF indicator.

Other noteworthy results involved median frequency, activation level, mobility, fractal Higuchi, spectral entropy and fuzzy entropy of the triceps that significantly changed for 75%, 60%, 67%, 85%, 70%, 70% of the participants respectively (Cohens’ *d* effect sizes ranged between 1.9-3.8); the activation level, activity, Higuchi fractal dimension, and spectral entropy of the biceps changed for 80%, 79%, 76%, 76%, (Cohens’ *d* effect sizes ranged between 2.26-2.83); the activation level and the activity of the upper trapezius that changed for 62% and 60 of the participants, respectively (Cohens’ *d* effect sizes equal to 0.67 and 0.63, respectively); the median frequency, the activation level and the mobility of the serratus anterior changed for 70% of the participants for all the three indicators (Cohens’ *d* effect sizes ranged between 1.5-1.8); the activation level and the Higuchi fractal dimension of the posterior deltoid changed for 75% and 70% of the participants (Cohens’ *d* effect sizes equal to 2.64 and 1.9, respectively); the degree of multifractality, multiscale entropy, percent of recurrence, correlation dimension, and largest Lyapunov exponent changed significantly for less than 45% of participants between RPT initiation and termination with Cohens’ *d* effect sizes lower than 0.7; the middle and lower trapezius showed significant changes for less than 57% of the participants (Cohen’s *d* effect sizes lower than 1.9) for all MMF indicators.

## 4 Discussion

The aim of this study was to identify the MMF indicators that best assess muscle fatigue during a repetitive pointing task. To this end, a PLSR method was used to predict the RPE from 15 linear and non-linear MMF indicators. The trained PLSR model explained 73% of the RPE variance. Median frequency, mobility, spectral entropy, fuzzy entropy, and fractal Higuchi had greater VIP values and changed for more than 65% of the participants during the RPT for both the anterior and medial deltoids. Moreover, the degree of multifractality, the multiscale entropy, the correlation dimension, the recurrence quantification analysis, and the largest Lyapunov exponent were the MMF indicators that had the lowest VIP values for the anterior and medial deltoids and did not change for a large proportion of participants.

### 4.1 Effect of RPT on muscle fatigue

As a decrease in MVIC and an increase in RPE is often used to emphasize muscle fatigue [1 - 4], the results of this study confirm the presence of muscular fatigue at RPT termination. Indeed, the t-test analysis revealed a significant decrease of the MVIC performed immediately after the completion of the RPT compared to the MVIC performed before the RPT. Also, all participants reached 8 or higher out of 10 at RPE scale at the completion of the RPT, 7 corresponding to “very strong” and 10 to “extremely strong (almost max)” perceived exertion [69] to perform the RPT at task termination. Since the task remained the same across the RPT, the increasing perceived exertion, in combination with the decreasing maximum voluntary isometric force indicated that the RPT caused muscle fatigue [107].

### 4.2 PLSR algorithm to predict RPE

For the first time, the present study used PLSR to predict muscle fatigue through the evolution of RPE and using MMF indicators as predictors during multi-joint movement of the shoulder. This approach allowed to explain 73% of the RPE variance on the training set, with an absolute error of prediction of 1.36 on the testing set. This explained variance is greater than that observed by Goubault et al., [24], Ni et al., [68] and Troiano [43] that used bivariate of multivariate regression models. Indeed, Goubault et al., [24] performed multivariate regression analyses between various MMF indicators and RPE scores during a working task. Their R-square values ranged between 11% and 39%. In the study of Troiano [43], the EMG fractal dimension and the EMG normalized mean frequency explained the RPE changes by 52% and 50%, respectively, during isometric contractions of the upper trapezius. In Ni et al., [68], EMG indicators explained 49% of the RPE variance during dynamic contractions. In the present study, the greater percentage of RPE variance explained can be due to the combination of several MMF indicators together in the PLSR model. In addition, various complex indicators identified as good predictors of muscle fatigue [28], such as fuzzy entropy and Higuchi fractal dimension, were used for the first time in such a model. Interestingly, our PLSR approach not only provided better prediction of RPE, but also physiologically meaningful results. Indeed, the highest VIP values were observed for the anterior and medial deltoids that are the muscles known to fatigue the most during the RPT used in the present study [72,73] and for the median frequency, a commonly well recognized indicator of muscle fatigue [108–113].

### 4.3 Median frequency, mobility, spectral entropy, fuzzy entropy, and fractal Higuchi MMF indicators to assess muscle fatigue

Median frequency was the MMF indicator that had the highest VIP value (2.28) and changed for a very large proportion of the participants (>85%) between RPT initiation and termination in the muscles known to fatigue the most during the RPT used in the present study, i.e., anterior and medial deltoids [72,73]. This result agrees with Goubault et al., [24], that also found that when correlating six MMF indicators to RPE, mobility and spectral entropy were among the MMF indicators that showed the highest R-square values. Our results are consistent with this recent study as mobility and spectral entropy also showed high VIP values (1.26-1.41) and significantly changed for a large proportion of the participants (80%-86%) between RPT initiation and termination for the anterior and medial deltoids, which reaffirms that these MMF indicators may be relevant to assess muscle fatigue. Concerning chaos, correlation, entropy, and fractals MMF indicators, Higuchi fractal dimension and fuzzy entropy were the only indicators that had high VIP values (1.26-2.03) and changed significantly for a large proportion of the participants (65%-90%) for the anterior and medial deltoids. Higuchi fractal dimension therefore confirms that the fractal dimension is sensitive to intrinsic changes of EMG signals occurring with muscle fatigue [18,49]. As to fuzzy entropy, our results are in line with Xie et al., [54] which stated that it provides an improved evaluation of time series complexity. Considering that median frequency is sensitive to both muscle fiber conduction velocity and motor unit synchronization [17,114], mobility is sensitive to conduction velocity [56], spectral entropy is sensitive to muscle fiber conduction velocity and motor units firing rate [56], and fuzzy entropy and Higuchi fractal dimension are sensitive to the increase of motor unit synchronization [90,104] occurring with muscle fatigue, our results emphasize the need to consider and combine a variety of MMF indicators that are sensitive to all parameters of motor unit behavior affected by fatigue in order to improve its assessment. Noteworthy, median frequency, mobility, spectral entropy, fuzzy entropy, and Higuchi fractal dimension were the only indicators that both showed low VIP values and changed for a moderate proportion of the participants (14%-38%) during RPT initiation and termination for the trapezius muscles, which did not show signs of fatigue during the RPT used in the present study [72–74]. These five indicators may therefore have good specificity to assess muscle fatigue, and future investigations should focus on their combination into a single MMF indicator to maximize the sensitivity and specificity of muscle fatigue assessment.

### 4.4 Activation level, activity, approximate, sample, and multiscale entropy, recurrence quantification analysis, correlation dimension, and chaos MMF indicators to assess muscle fatigue

The activation level was shown to be a part of the two MMF indicators required to assess muscle fatigue according to the JASA method [14]. Firstly, our results showed that the VIP value of activation level of the medial deltoid was 1.41, which was significantly lower than the VIP value of the median frequency, mobility, Higuchi fractal dimension, spectral entropy, and fuzzy entropy for this muscle. Secondly, although the activation level of both the anterior and medial deltoid increased significantly for a large proportion of the participants (76%-80%), which can be interpreted as a MMF [14], the activation level of the posterior deltoid and upper trapezius also increased for a comparable proportion of the participants (75%-76%) between the RPT initiation and termination while they the latter muscles did not show sign of fatigue in previous studies [72,73]. Supporting these results on activation level, muscles act in synergy so that many postural and movement adaptations are induced by muscle fatigue during a RPT [72]. Therefore, the upper trapezius and the posterior deltoid changes in amplitude-based MMF indicators may reflect compensatory mechanisms in response to fatigue in prime mover muscles rather than local fatigue [115,116]. Consequently, a change of activation level may reflect local muscle fatigue for prime mover muscles or compensatory mechanisms in response to fatigue of prime mover muscles for synergistic muscles [72,73]. These observations suggest that activation level, and therefore the JASA method, should be used with caution for the assessment of muscle fatigue [6] as it may increase the risk of false positive. Another MMF indicator related to the amplitude of EMG signals is the activity. The VIP value of the activity of the median and anterior deltoids ranged between 0.73-1.61, which is qualitatively smaller than the VIP value of median frequency, mobility, Higuchi fractal dimension, spectral entropy, and fuzzy entropy (1.26-2.28) for these muscles. Similarly to activation level, there were changes in activity for a large proportion of the participants in the upper trapezius (90%), although this muscle is not supposed to fatigue [72,73]. Thus, our results further confirm that amplitude related MMF indicators are not relevant for assessing muscle fatigue.

Approximate and sample entropy had moderate VIP values (0.91-1.57), in the anterior and median deltoids and changed for a moderate to large proportion of the participants (52%-76%) between the RPT initiation and termination. To support this latter result, the lack of consistency and monotonicity of approximate entropy causes difficulty in interpreting the signal’s complexity and reduced its efficiency to dichotomize fatigue and non-fatigue states [54,88]. Although sample entropy addresses the drawbacks of approximate entropy calculation to assess muscle fatigue [88], our results revealed that this MMF indicator had moderate efficiency to assess muscle fatigue, which may be caused by its high variability to precise parameter selection [117]. Multiscale entropy was therefore introduced to better detect the presence of complexity in time series and overcome the limitations of approximate and sample entropy [57]. Although multiscale entropy was shown more sensitive to muscle fatigue than median frequency [118], our results evidenced that multiscale entropy had significantly smaller VIP values than median frequency and changed for a moderate proportion of the participants (35%-45%) for the anterior and medial deltoids between RPT initiation and termination. Consequently, fuzzy entropy, as previously discussed, is the only entropy-based indicator that may be efficient to assess MMF. Concerning the degree of multifractality, it had very low VIP values (<0.5) and marginally changed (<10%) between the RPT initiation and termination. This result contradicts previous studies that found that this multifractal MMF indicator is more efficient than the median frequency and Higuchi fractal dimension to assess muscle fatigue in dynamic and static conditions [40,46,47,49]. More investigations are therefore required to determine the effectiveness of the degree of multifractality for assessing MMF. As to correlation dimension, percent determinism, and percent of recurrence, they had very low VIP values (0.4-1.2), which agrees with previous evidences that correlation dimension is not sensitive to changes in EMG signal properties caused by fatigue [28]. Although previous studies have shown an effect of muscle fatigue on recurrence quantification analyses, our results are in agreement with the lower performance of this MMF indicator compared to frequency-based and fuzzy entropy indicators [75]. Finally, the largest Lyapunov exponent also had very low VIP values for all muscles (<0.6) and did not change during the RPT for more than 38% of the participants between RPT initiation and termination in median and anterior deltoid. To support this result, Chakraborty et al., [36] found that the largest Lyapunov exponent did not change with fatigue during dynamic contractions of the biceps brachii. Thus, our results confirm that the potential of approximate, sample, and multiscale entropy, degree of multifractality, correlation dimension, and the largest Lyapunov exponent is limited to assess muscle fatigue during low-load repetitive tasks, which question their sensitivity to changes in muscle fiber conduction velocity [5,84,104] and motor unit firing rate [35,56,103] and synchronization [35,104] caused by fatigue.

### 4.5 Limitations

The first limitation of this study was that, considering the nature of the task which is a multi-joint task performed with the whole dominant upper limb in a standing position, several muscles act as prime mover and stabilizer muscles so that changes of MMF indicators may be caused either by their function or muscle fatigue that make interpretation of amplitude related indicators difficult [115,119]. Another limitation is that the RPE was used as a measure of fatigue, while its evolution can be multifactorial during a fatiguing task. Finally, even though the previous literature has suggested the presence of sex differences in some EMG patterns of fatigue during the performance of the RPT [120], our sample size was too small to develop a sex-specific model.

## 5 Conclusion

The present study confirmed that median frequency, mobility, and spectral entropy are efficient EMG-based indicators to assess muscle fatigue. Interestingly, our analyses also showed that fuzzy entropy and Higuchi fractal dimension may also be efficient MMF indicators. Therefore, a combination of median frequency, mobility, spectral entropy, fuzzy entropy, and Higuchi fractal dimension should be further considered to assess muscle fatigue compared to the exclusive use of median frequency and activation level. Alternatively, although other MMF indicators such as the multiscale entropy, the degree of multifractality, correlation dimension, the percent of determinism, the percent of recurrence, and the largest Lyapunov exponent were previously used to assess muscle fatigue, they should no further be considered as our analyses showed that they poorly contributed to the prediction of RPE and did not change significantly for a large proportion of the participants during a fatiguing RPT. These results may help to improve the identification of MMF indicators to accurately assess muscle fatigue. In the long term, more accurate methods to identify fatigue-related changes in EMG, combined with other advances such as in workplace wearable sensors, could contribute to the real-time, early detection of injury risk factors, and ultimately, prevention of musculoskeletal disorders caused by muscle fatigue.

## Data Availability

All data produced in the present study are available upon reasonable request to the authors

## 6 Acknowledgments

We would like to thank our industrial partner, David’s TEA, with a special mention to our contact, R. Butman, for his help in the recruitment.

