## Supplementary file for "What are the best indicators of myoelectric manifestation of fatigue?"

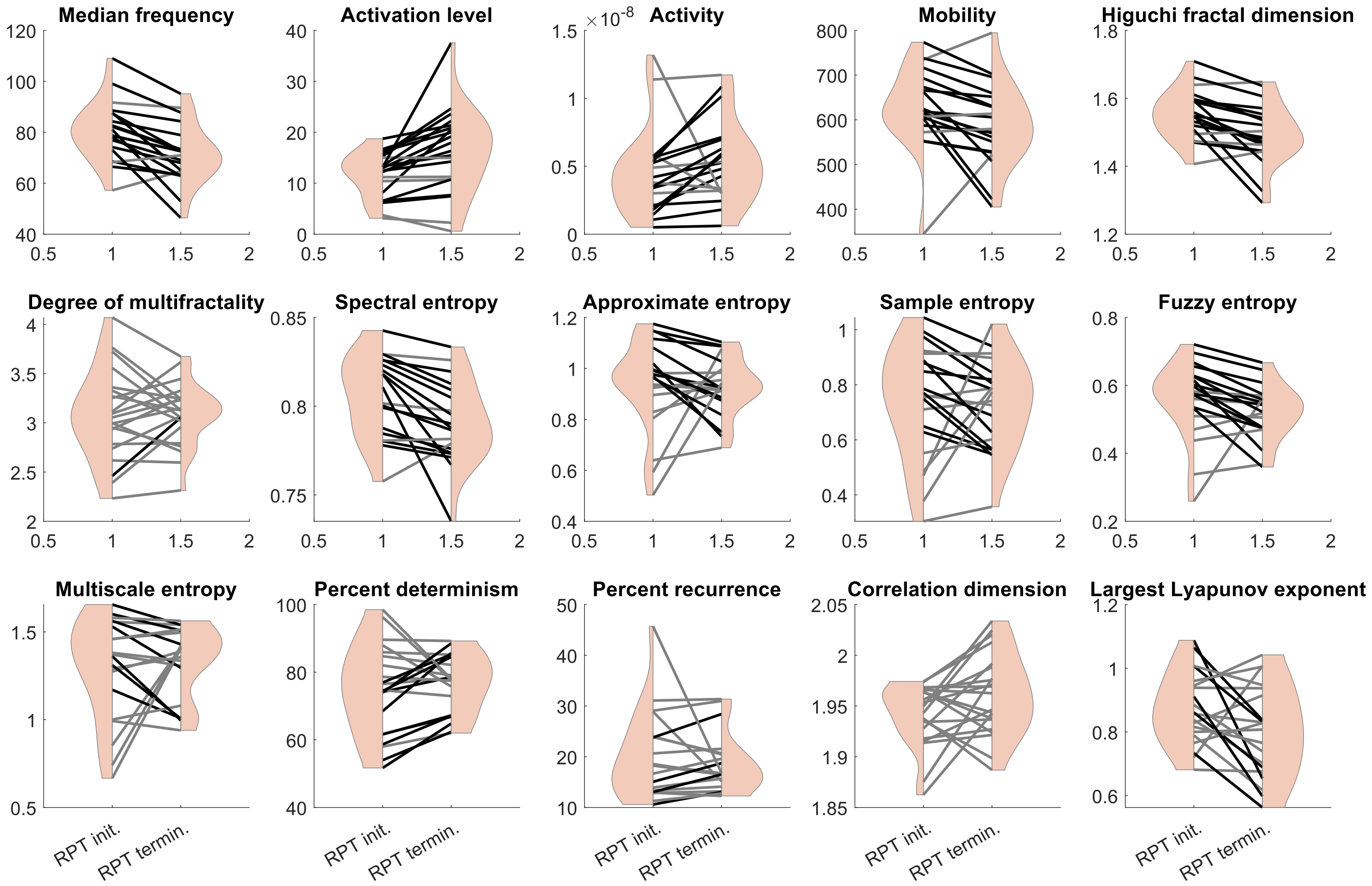


Figure S1: MMF indicators for the anterior deltoid at RPT initiation and termination. Each line represents data from one participant. Black lines indicate a significant difference between RPT initiation and termination in the direction, i.e., increase or decrease, reported by the literature in presence of muscle fatigue. Grey lines indicate no difference between RPT initiation and termination.

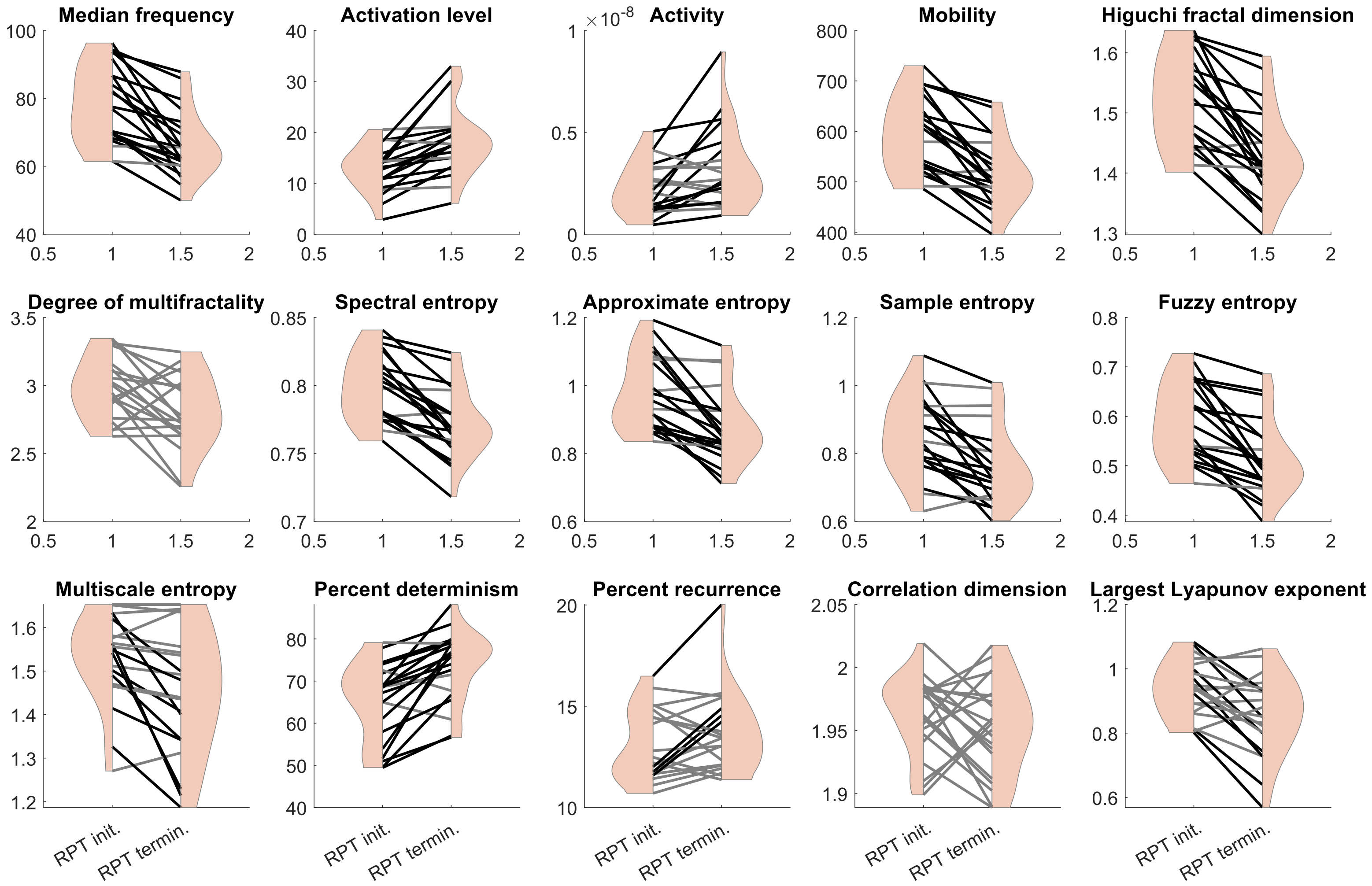


Figure S2: MMF indicators for the medial deltoid at RPT initiation and termination. Each line represents data from one participant. Black lines indicate a significant difference between RPT initiation and termination in the direction, i.e., increase or decrease, reported by the literature in presence of muscle fatigue. Grey lines indicate no difference between RPT initiation and termination.

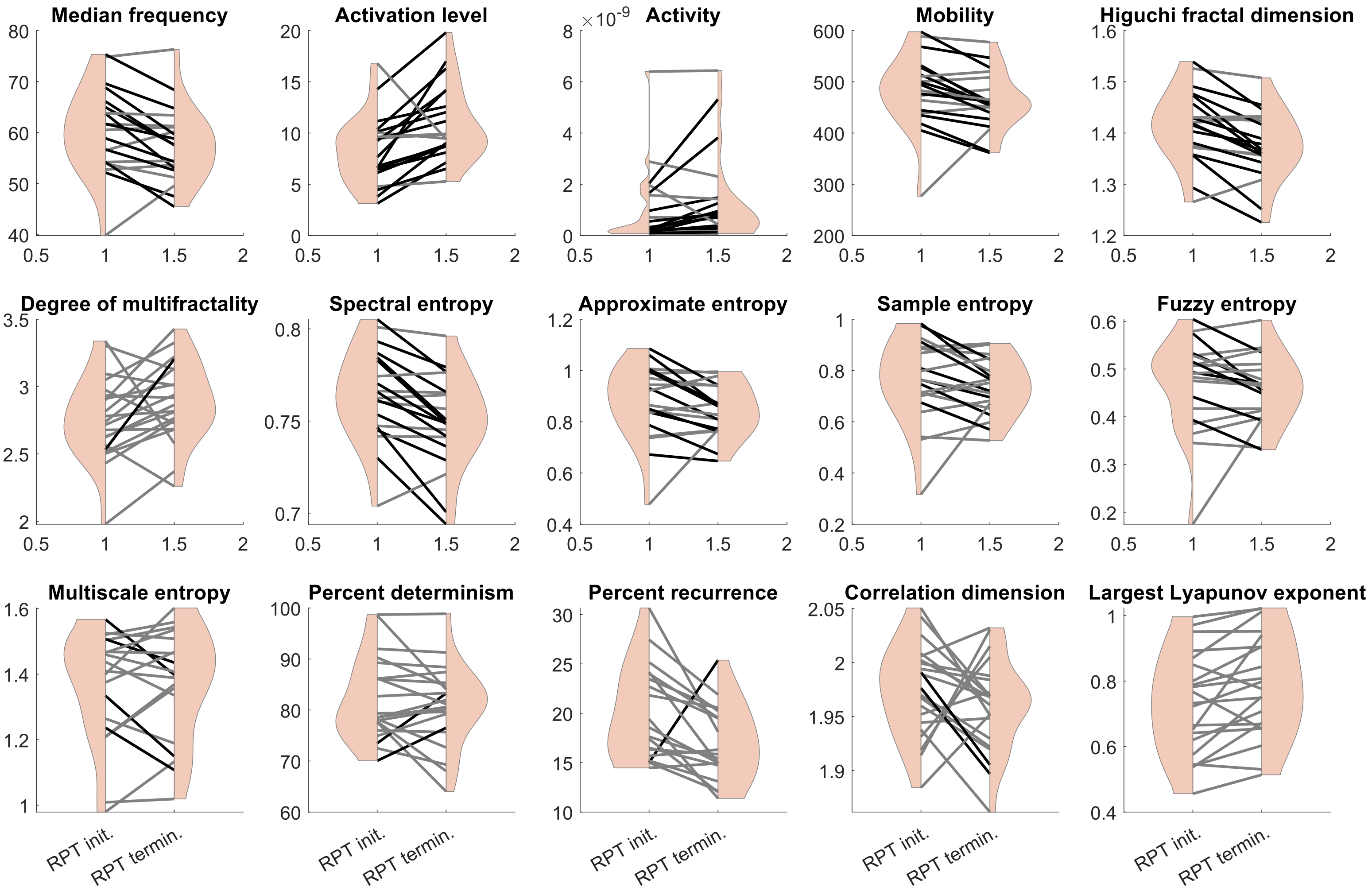


Figure S3: MMF indicators for the posterior deltoid at RPT initiation and termination. Each line represents data from one participant. Black lines indicate a significant difference between RPT initiation and termination in the direction, i.e., increase or decrease, reported by the literature in presence of muscle fatigue. Grey lines indicate no difference between RPT initiation and termination.

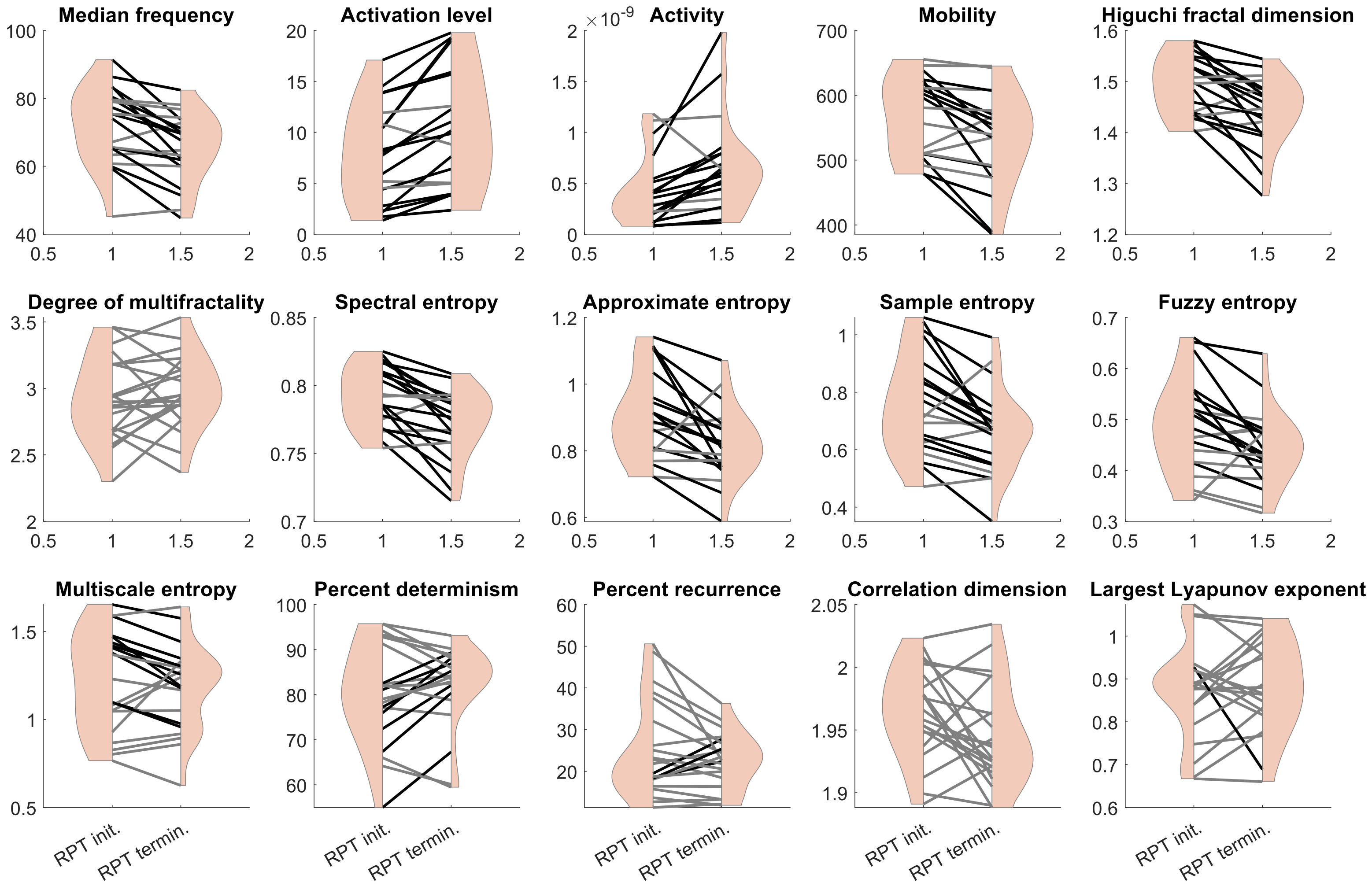


Figure S4: MMF indicators for the biceps at RPT initiation and termination. Each line represents data from one participant. Black lines indicate a significant difference between RPT initiation and termination in the direction, i.e., increase or decrease, reported by the literature in presence of muscle fatigue. Grey lines indicate no difference between RPT initiation and termination.

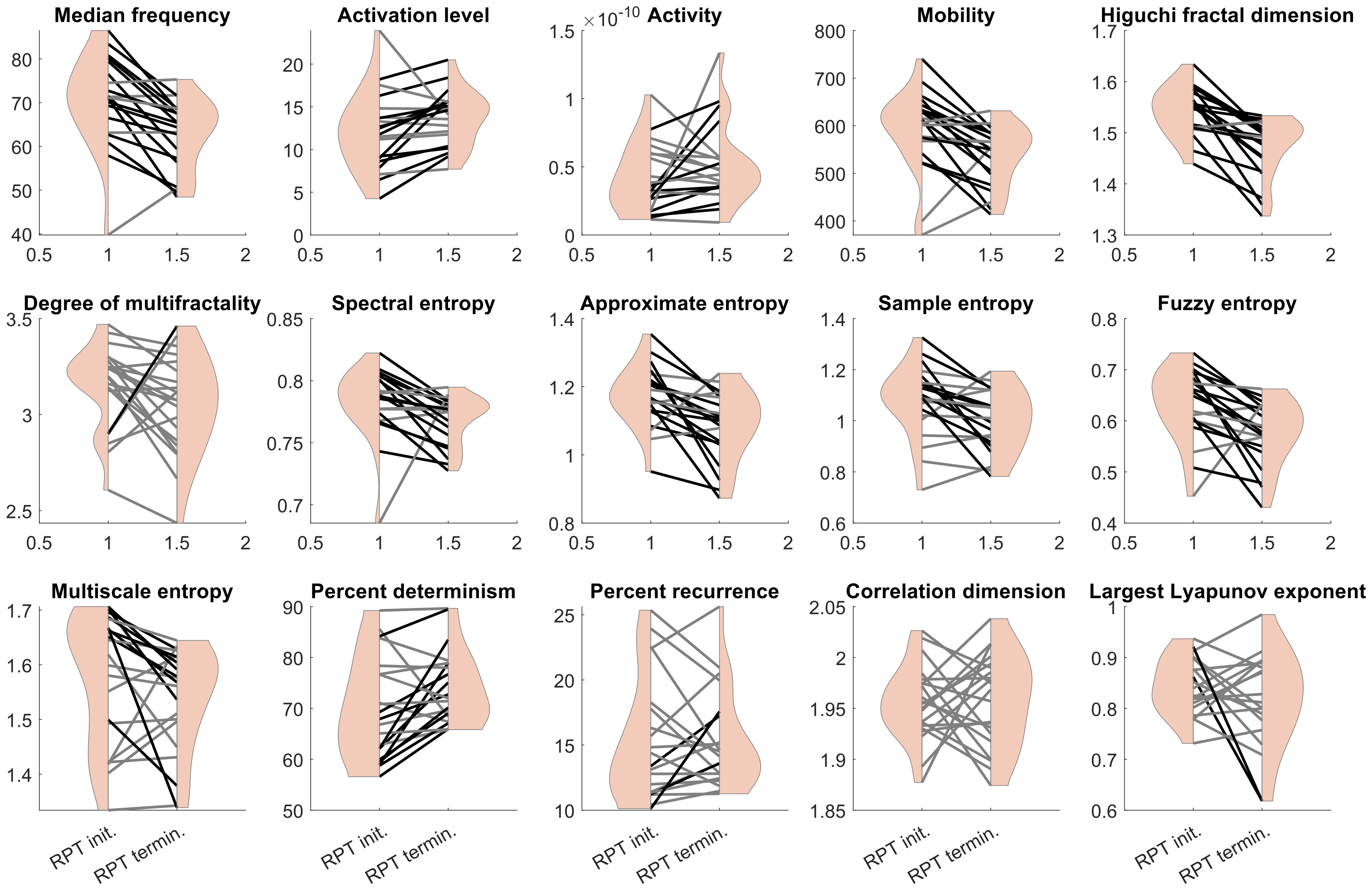


Figure S5: MMF indicators for the triceps at RPT initiation and termination. Each line represents data from one participant. Black lines indicate a significant difference between RPT initiation and termination in the direction, i.e., increase or decrease, reported by the literature in presence of muscle fatigue. Grey lines indicate no difference between RPT initiation and termination.

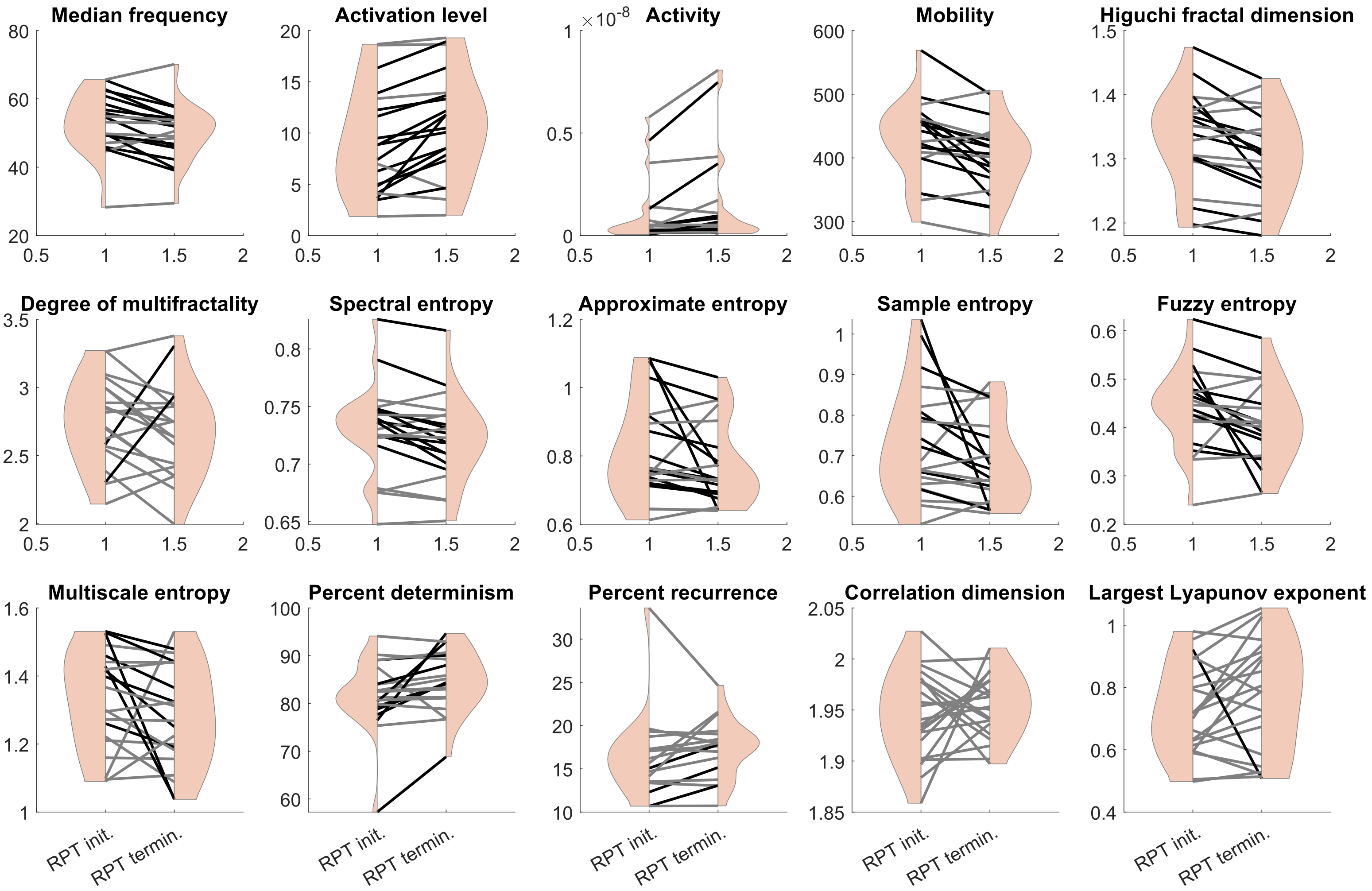


Figure S6: MMF indicators for the serratus anterior at RPT initiation and termination. Each line represents data from one participant. Black lines indicate a significant difference between RPT initiation and termination in the direction, i.e., increase or decrease, reported by the literature in presence of muscle fatigue. Grey lines indicate no difference between RPT initiation and termination.

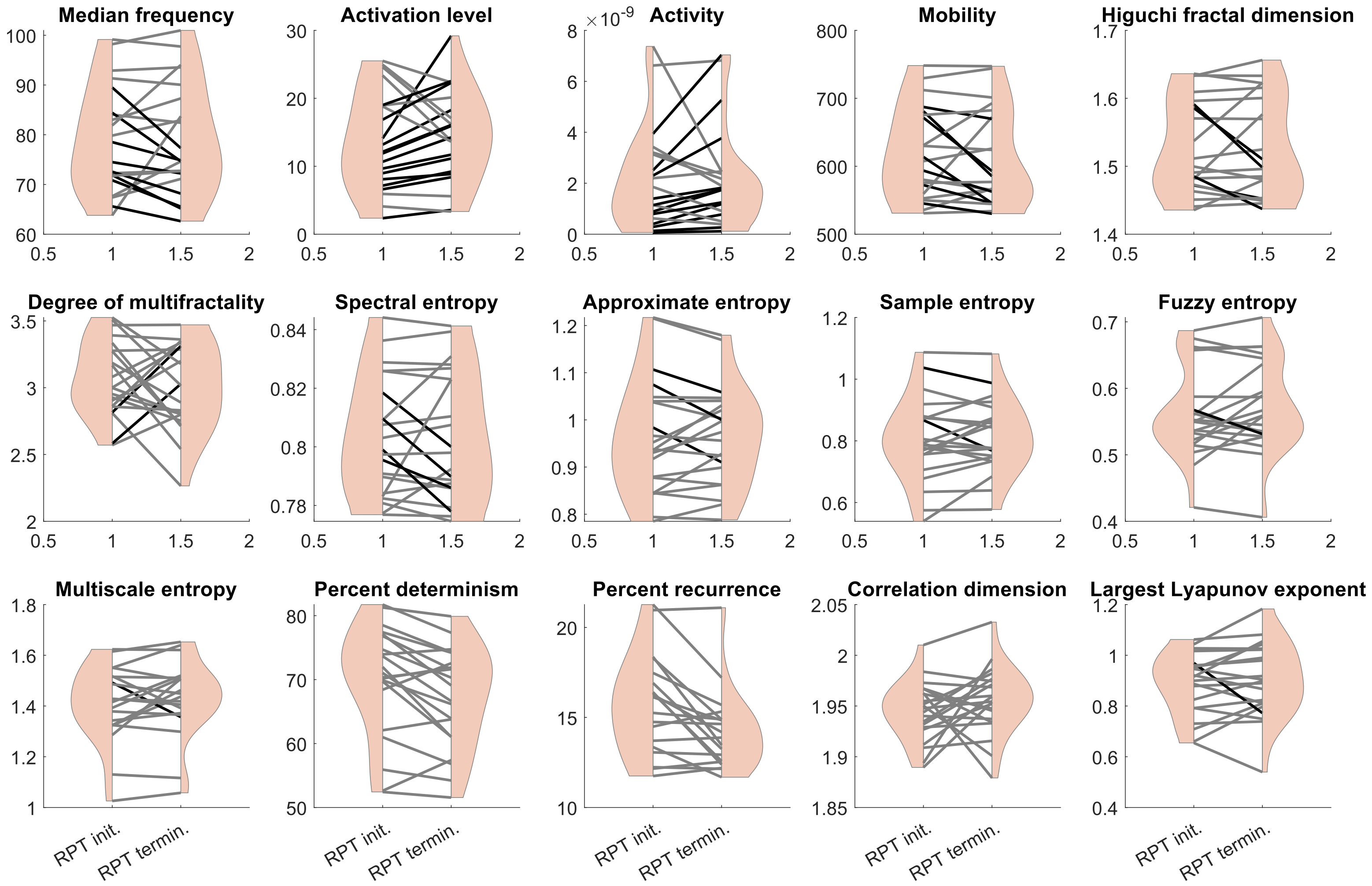


Figure S7: MMF indicators for the upper trapezius at RPT initiation and termination. Each line represents data from one participant. Black lines indicate a significant difference between RPT initiation and termination in the direction, i.e., increase or decrease, reported by the literature in presence of muscle fatigue. Grey lines indicate no difference between RPT initiation and termination.

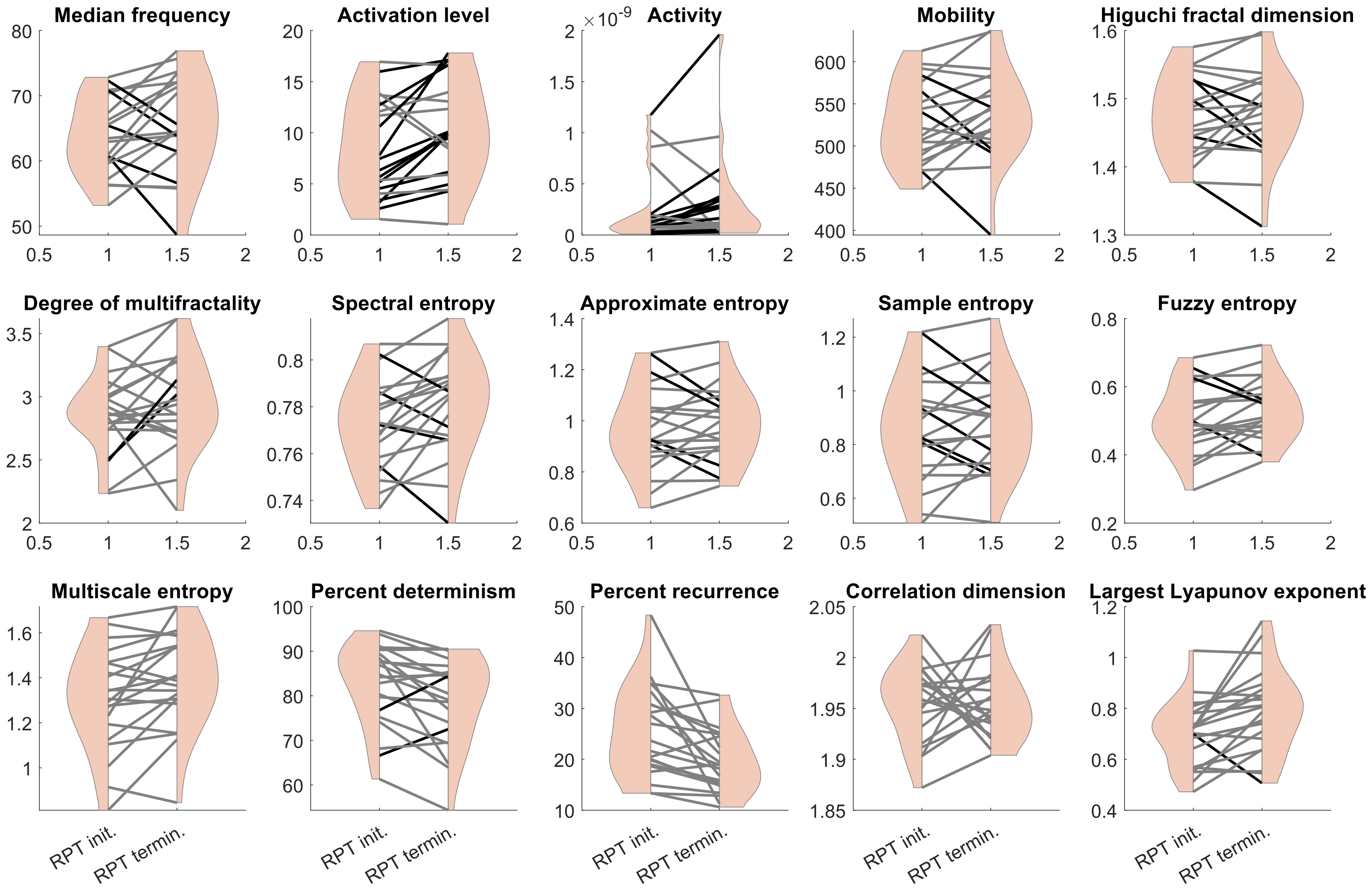


Figure S8: MMF indicators for the middle trapezius at RPT initiation and termination. Each line represents data from one participant. Black lines indicate a significant difference between RPT initiation and termination in the direction, i.e., increase or decrease, reported by the literature in presence of muscle fatigue. Grey lines indicate no difference between RPT initiation and termination.

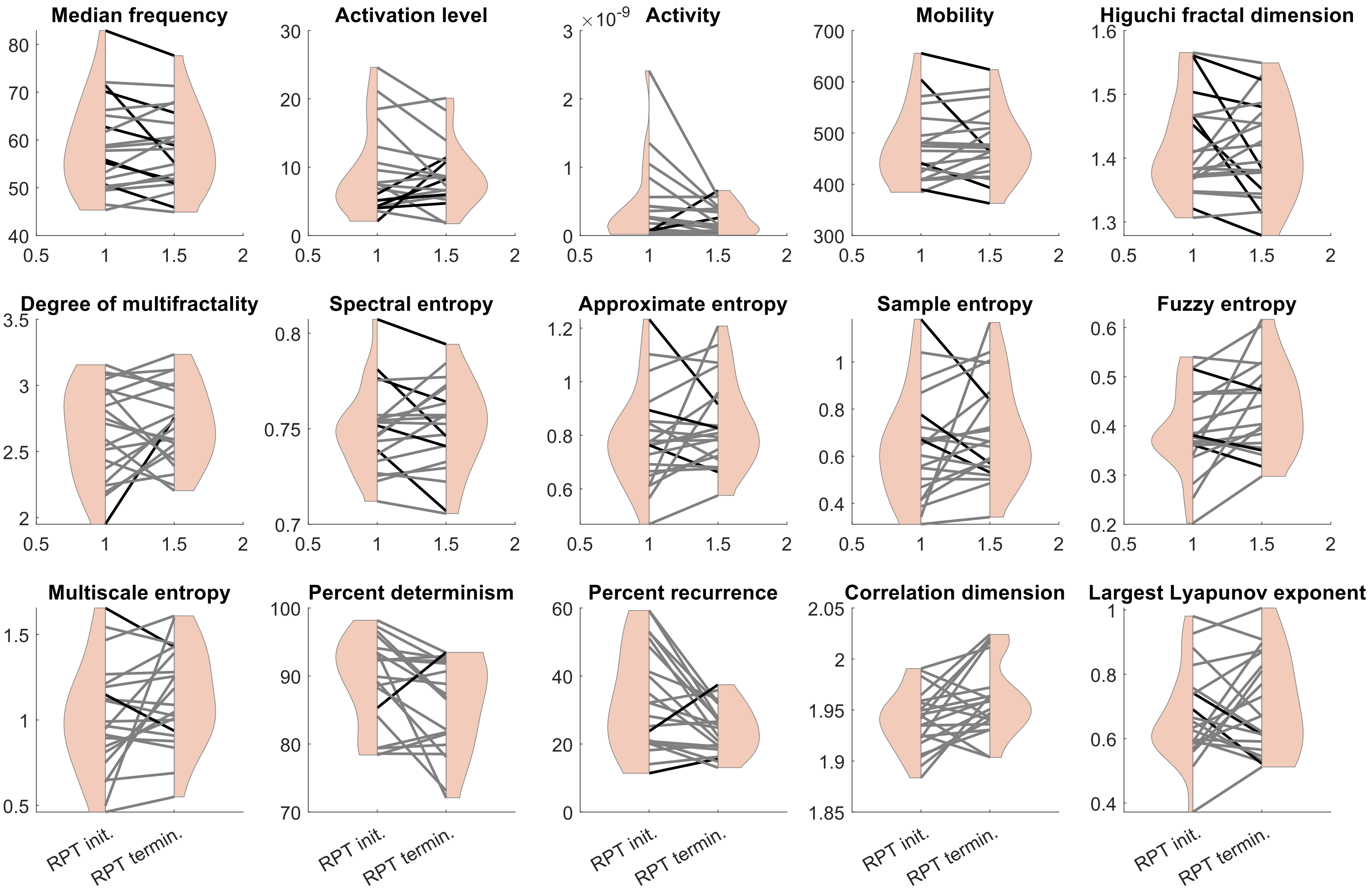


Figure S9: MMF indicators for the lower trapezius at RPT initiation and termination. Each line represents data from one participant. Black lines indicate a significant difference between RPT initiation and termination in the direction, i.e., increase or decrease, reported by the literature in presence of muscle fatigue. Grey lines indicate no difference between RPT initiation and termination.
